# Who has long-COVID? A big data approach

**DOI:** 10.1101/2021.10.18.21265168

**Authors:** Emily R. Pfaff, Andrew T Girvin, Tellen D. Bennett, Abhishek Bhatia, Ian M. Brooks, Rachel R Deer, Jonathan P Dekermanjian, Sarah Elizabeth Jolley, Michael G. Kahn, Kristin Kostka, Julie A McMurry, Richard Moffitt, Anita Walden, Christopher G Chute, Melissa A Haendel

**Affiliations:** UNC Chapel Hill School of Medicine, Department of Medicine, 160 N Medical Drive, Chapel Hill, NC 27599; Palantir Technologies, Denver, CO; Sections of Informatics and Data Science and Critical Care Medicine, Department of Pediatrics, University of Colorado School of Medicine, Aurora, CO; Carolina Health Informatics Program, University of North Carolina at Chapel Hill, Chapel Hill, NC; Colorado Center for Personalised Medicine, Division of Biomedical Informatics & Personalized Medicine, Department of Medicine, University of Colorado Anschutz Medical Center; Department of Nutrition, Metabolism, and Rehabilitation Sciences, University of Texas Medical Branch, Galveston, TX USA 77550; Department of Biostatistics and Informatics, Colorado School of Public Health, University of Colorado-Denver Anschutz Medical Campus; Department of Medicine, Division of Pulmonary and Critical Care Medicine, University of Colorado Anschutz Medical Campus, Aurora, CO 80045; Section of Informatics and Data Science, Department of Pediatrics, University of Colorado School of Medicine, Aurora, CO; The OHDSI Center at the Roux Institute, Northeastern University, Portland, ME; Center for Health AI, University of Colorado Anschutz Medical Campus, Aurora, CO; Department of Biomedical Informatics, Stony Brook Cancer Center, Stony Brook University, Stony Brook, NY; Schools of Medicine, Public Health, and Nursing, Johns Hopkins University, Baltimore, MD; University of Colorado Anschutz Medical Campus, Aurora, CO

## Abstract

**Background:** Post-acute sequelae of SARS-CoV-2 infection (PASC), otherwise known as long-COVID, have severely impacted recovery from the pandemic for patients and society alike. This new disease is characterized by evolving, heterogeneous symptoms, making it challenging to derive an unambiguous long-COVID definition. Electronic health record (EHR) studies are a critical element of the NIH Researching COVID to Enhance Recovery (RECOVER) Initiative, which is addressing the urgent need to understand PASC, accurately identify who has PASC, and identify treatments.

**Methods:** Using the National COVID Cohort Collaborative’s (N3C) EHR repository, we developed XGBoost machine learning (ML) models to identify potential long-COVID patients. We examined demographics, healthcare utilization, diagnoses, and medications for 97,995 adult COVID-19 patients. We used these features and 597 long-COVID clinic patients to train three ML models to identify potential long-COVID patients among (1) all COVID-19 patients, (2) patients hospitalized with COVID-19, and (3) patients who had COVID-19 but were not hospitalized.

**Findings:** Our models identified potential long-COVID patients with high accuracy, achieving areas under the receiver operator characteristic curve of 0.91 (all patients), 0.90 (hospitalized); and 0.85 (non-hospitalized). Important features include rate of healthcare utilization, patient age, dyspnea, and other diagnosis and medication information available within the EHR. Applying the “all patients” model to the larger N3C cohort identified 100,263 potential long-COVID patients.

**Interpretation:** Patients flagged by our models can be interpreted as “patients likely to be referred to or seek care at a long-COVID specialty clinic,” an essential proxy for long-COVID diagnosis in the current absence of a definition. We also achieve the urgent goal of identifying potential long-COVID patients for clinical trials. As more data sources are identified, the models can be retrained and tuned based on study needs.

**Funding:** This study was funded by NCATS and NIH through the RECOVER Initiative.

## Background

Acute COVID-19 affects multiple organ systems, including the lungs, digestive tract, kidneys, heart, and brain.^1,2^ The longer term clinical consequences of COVID-19 are still poorly understood and are collectively termed post-acute sequelae of SARS-CoV-2 infection (PASC), or long-COVID.^3^ At this time, this disease is referred to by a number of terms that may or may not represent the same constellation of signs and symptoms; here, we consider PASC synonymous with long-COVID. Long-COVID can be broadly defined as persistent or new symptoms more than four weeks after severe, mild, or asymptomatic SARS-CoV-2 infection.^4,5^ Characterizing, diagnosing, treating, and caring for long-COVID patients has proven very challenging due to heterogeneous signs and symptoms that evolve over long trajectories.^6^ The impact of long-COVID on patients’ quality of life and ability to work can be profound.

The wide range of symptoms attributed to long-COVID was highlighted in an extensive patient-led survey,^7^ which conducted deep longitudinal characterization of long-COVID symptoms and trajectories in suspected and confirmed COVID-19 patients reporting illness lasting more than 28 days.^8^ Evaluation and harmonization of patient- and clinically reported long-COVID features using the Human Phenotype Ontology (HPO) also revealed heterogeneous signs and symptoms, supporting the conclusion that a complex constellation of both patient- and clinically reported features is necessary to correctly classify and manage long-COVID patients.^9^ The World Health Organization (WHO) recently published its own case definition of “post COVID-19 condition” (WHO’s term) that includes twelve criteria, which similarly require a wide variety of patient-declared and clinical information.^10^

In order to gain an understanding of the complexities of long-COVID, it will be necessary to recruit a large and diverse cohort of research participants. The NIH’s RECOVER study^11^ is one such initiative aiming to recruit thousands of participants nationwide in order to answer critical research questions about PASC such as understanding pregnancy risk factors, cognitive impairment and mental health, and outcome disparities and comorbidities. Efficient recruitment of cohorts of this size and scope often entails leveraging computable phenotypes^12–14^ (i.e., electronic cohort definitions) to find sufficient numbers of patients meeting a study’s inclusion criteria. Poor cohort definition can result in poor study outcomes.^15,16^ For long-COVID, as with other novel conditions, the lack of an agreed-upon definition and the heterogeneity of the condition’s presentation poses a significant challenge to cohort identification. Machine learning (ML) can help address this by using the rich longitudinal data available in electronic health records (EHRs) to algorithmically identify patients similar to those in a long-COVID “gold standard.”

The National COVID Cohort Collaborative (N3C)^17^ offers a data-driven solution to quantifying the features of long-COVID and an appropriate proving ground for an ML approach.^18^ N3C is an NIH National Center for Advancing Translational Sciences (NCATS)-sponsored data and analytic environment which compiles and harmonizes longitudinal electronic EHR data from 65 sites and over 8 million patients who have (1) tested positive for SARS-CoV-2 infection, (2) whose symptoms are consistent with a COVID-19 diagnosis, or (3) are demographically matched controls who have tested negative for SARS-CoV-2 infection (and have never tested positive) to support comparative studies.^19^ To build a foundation for a robust clinical definition of long-COVID, we linked curated long-COVID clinic patient lists from three N3C sites with data in the N3C repository. We then used the linked dataset to train and test three ML models and applied those models to (1) define a nationwide cohort of potential long-COVID patients and (2) derive a list of prominent clinical features shared among that cohort to help identify patients for research studies and target features for further investigation.

## Research in context

### Evidence before this study

Initial characterization of long-COVID patients has contributed to an emerging clinical understanding, but the significant heterogeneity of disease features makes diagnosing and treating this new disease extremely challenging. This challenge is urgent to address, as many patients report long-COVID symptoms as debilitating, significantly affecting their ability to engage in activities of daily life. Few studies have utilized large-scale databases to understand concordance of clinical patterns and generate data-driven definitions of long-COVID. The NIH RECOVER program has invested in EHR studies to understand PASC, to accurately identify who has PASC, and to prevent and treat PASC.

### Added value of this study

N3C harmonizes patient-level EHR data from over 8 million demographically diverse and geographically distributed patients. Here, we describe highly accurate XGBoost ML models that use N3C to identify presumptive long-COVID patients, trained using EHR data from patients who attended a long-COVID specialty clinic. The most powerful predictors in these models are outpatient clinic utilization post-acute COVID-19, patient age, dyspnea, and other diagnosis and medication features readily available in the EHR. The model is transparent and reproducible, and can be widely deployed at individual healthcare systems to enable local research recruitment or secondary data analysis.

### Implications of all the available evidence

N3C’s longitudinal data for COVID-19 patients provides a comprehensive foundation for the development of ML models to identify potential long-COVID patients. Such models enable efficient study recruitment efforts to deepen our understanding of long-COVID and offer opportunities for hypothesis generation. Moreover, as more patients are diagnosed with long-COVID and novel sources of data become more available (e.g., sensor data from wearables), our models can be refined and retrained, evolving the algorithm as more evidence emerges. Our model can also be the basis for predicting who will seek care for long-COVID, and therefore provide the basis for generalizable clinical decision support tools.

## Methods

### Selecting and subsetting the base population

To model long-COVID, we used EHR data integrated and harmonized in Palantir Foundry inside the N3C Secure Data Enclave to identify unique healthcare utilization patterns and clinical features among COVID-19 patients. For the purpose of this study, we defined our base population (*n* = 1,793,604) as any non-deceased adult patient (age >= 18 years) with either an ICD-10-CM COVID-19 diagnosis code (U07.1) from an inpatient or emergency visit, *or* a positive SARS-CoV-2 PCR or antigen test, and for whom at least 90 days have passed since COVID-19 index date. Prior to this analysis, patients from six N3C sites were removed from the cohort due to their sites’ use of randomly shifted dates of service, which limits our ability to use temporal logic during analysis.

Without an agreed-upon case definition, no long-COVID gold standard exists to validate computable phenotypes and train ML models. However, three N3C sites provided lists of locally identified patients who had visited that site’s long-COVID speciality clinic one or more times. These patients represent a “silver standard” within our base population (*n =* 597, once our base population criteria were applied). Hereafter, we will refer to this group of patients as “long-COVID clinic patients”; patients identified by our trained model as long-COVID patients will be referred to as “potential long-COVID patients.” This silver standard enabled us to develop a model to identify patients likely to be referred to or seek care at a long-COVID clinic—a valuable proxy for long-COVID until a true gold standard is available.

For the purposes of training and testing ML models, we created a subset of our overall cohort containing only patients originating from those three sites (*n* = 97,995), including the long-COVID clinic patients. This subset was stratified further into patients who had been hospitalized with acute COVID-19 (*n =* 19,368) and patients not hospitalized (*n =* 78,627). We narrowed further to patients who had at least one encounter and at least one diagnosis *or* one medication in their post-COVID-19 window (15,621 hospitalized, 58,351 not hospitalized). The full cohort selection and subsetting process is illustrated in Supplemental Figure 1.

### Feature selection

Within the three-site subset, we examined demographics, encounter details, conditions, and medication orders for each patient before and after their period of acute COVID-19. Though its use was considered, lab result data ultimately proved too sparse among the cohort for use in the models, especially for non-hospitalized patients. Features were selected for inclusion in the model by gathering data points in these domains associated with the long-COVID clinic patients in the time period of interest (see Figure 1). For each patient, we only counted diagnoses that newly occurred or occurred in greater frequency in the post-COVID-19 period compared to the pre-COVID-19 period, and only counted medications that were newly prescribed in the post-COVID-19 period, with no order records in the pre-COVID-19 period. Detailed feature engineering methods are described in Supplemental Methods.

**Figure 1.**
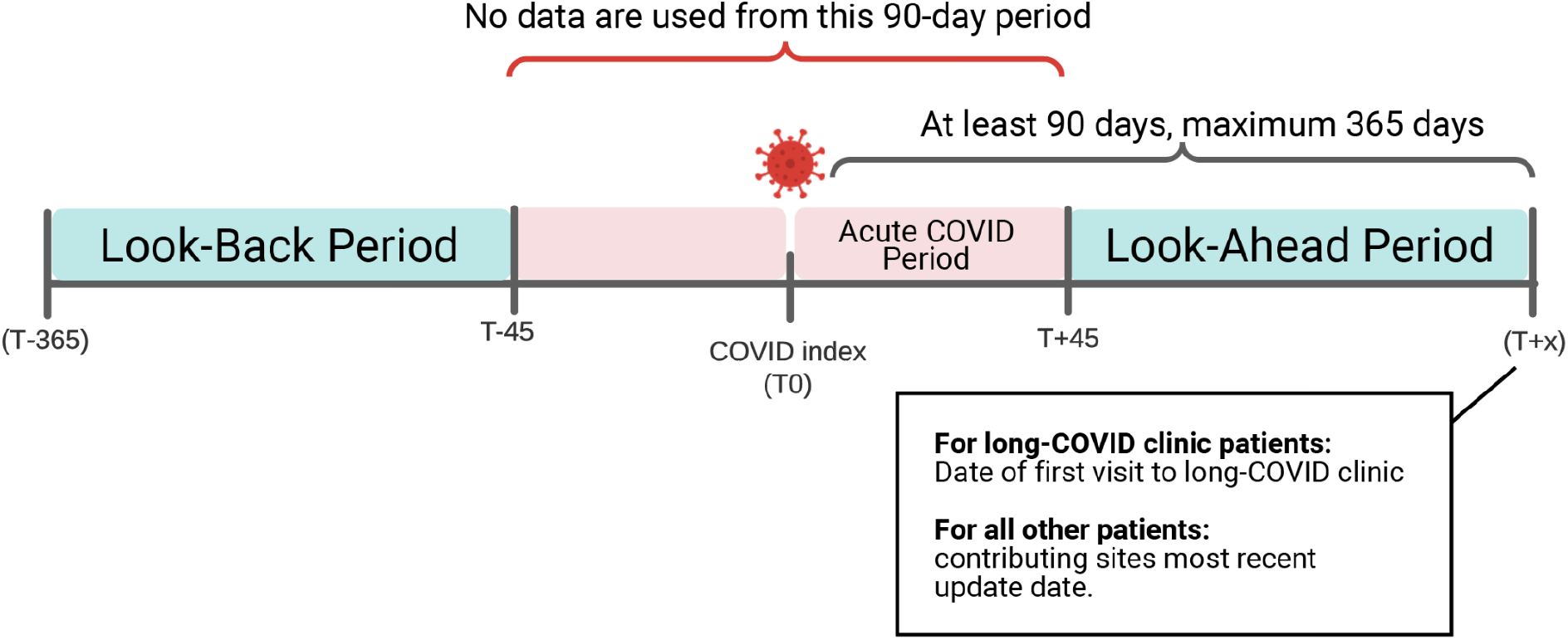
Temporal windows for ML model inclusion. We searched for encounters, conditions, and medication orders before and after each patient’s COVID-19 index, up to a maximum of 365 days post-index. We ignored all data occurring in a “buffer” period of 45 days before and after the COVID-19 index date to differentiate pre- and post-COVID-19 from acute COVID-19. For long-COVID clinic patients, we ignored all data occurring on or after their first visit to a long-COVID clinic to avoid influencing the model with clinical observations occurring as a result of the patient’s long-COVID assessment.

### Modeling

To reflect that long-COVID may look different depending on the severity of the patient’s acute COVID-19, we built three different ML models using the three-site subset: (1) all patients, (2) patients who had been hospitalized with acute COVID-19, and (3) patients who were not hospitalized. The intent of each model is to identify the patients most likely to have long-COVID, using attendance at a long-COVID specialty clinic as a proxy for long-COVID diagnosis. To train and test each model, patients were randomly sampled to yield similar patient counts in both classes (long-COVID clinic patients and patients who did not attend the long-COVID clinic). For the all-patient model, data were also sampled to yield similar numbers of hospitalized and non-hospitalized patients. Counts of patients in each group used for training and testing are shown in Supplemental Table 1.

The python package XGBoost was used to construct the models, using 924 features in total. Categorical features were one-hot encoded. Age and encounter rates were treated as continuous variables and conditions and drugs were modeled as binary features; see Supplemental Methods for feature engineering details. Model hyperparameters were tuned using scikit-learn’s GridSearchCV, with 5-fold cross validation, set to optimize the area under the receiver operating characteristic curve (AUROC). We trained each model using 5-fold cross validation, repeated 5 times. To assess performance, we calculated the AUROC, as well as the precision, recall, and F-score for each model with a predictive probability threshold of 0.45. To aid with interpretability, we calculated SHapley Additive exPlanation (SHAP) values^20^ for all features to quantify each feature’s importance to the classifications made by each model.

Once trained, we ran the “all patients” model over the full base population of patients who had at least one encounter and at least one diagnosis *or* one medication in their post-COVID-19 window (*n* = 846,981), in order to flag potential long-COVID patients in the N3C Enclave.

## Findings

The combined demographics of the long-COVID clinic patients supplied by the three sites (first two columns of Table 1) show significant differences from the COVID-19 patients at those sites who did not attend the long-COVID clinic (third and fourth columns of Table 1). Notably, non-hospitalized long-COVID clinic patients are disproportionately female. Long-COVID clinic patients who were hospitalized with acute COVID-19 are disproportionately Black when compared with all patients hospitalized with acute COVID-19, and are much more likely to have a pre-COVID-19 comorbidity (diabetes, kidney disease, congestive heart failure, or pulmonary disease).

**Table 1.** Characteristics of the three-site cohort used for model training and testing. All patients shown had acute COVID-19. In accordance with the N3C download policy,^21^ for demographics where small cell sizes (<20 patients) could be derived from context, we have shifted the counts +/- by a random number between 1 and 5. The accompanying percentages reflect the shifted number. All shifted counts are labelled as such, e.g. +/- 5.

|  | Long-COVID clinic |  | Not long-COVID clinic |  |
| --- | --- | --- | --- | --- |
|  | Hospitalized<br>n = 428 | Not hospitalized<br>n = 169 | Hospitalized<br>n = 15,193 | Not hospitalized<br>n = 58,182 |
| **Sex (%)** |  |  |  |  |
| Female | 237 (55.4) | 127 (75.1) | 8,465 +/-5 (55.7) | 34,771 (59.8) |
| Male | 191 (44.6) | 42 (24.9) | 6,716 +/-5 (44.2) | 23,258 (40.0) |
| Unknown | 0 (0.0) | 0 (0.0) | <20 | 153 (0.3) |
| **Age (mean (SD))** | 58.29 y (15.03) | 48.14 y (14.02) | 56.50 y (18.60) | 45.92 y (17.32) |
| **Race (%)** |  |  |  |  |
| Asian | <20 | <20 | 571 (3.8) | 1361 (2.3) |
| Black | 190 (44.4) | 31 (18.3) | 3,207 (21.1) | 6,370 (10.9) |
| Native Haw./Pac. Islander | <20 | <20 | 43 (0.3) | 138 (0.2) |
| Other | <20 | <20 | 85 (0.6) | 254 (0.4) |
| Unknown | 81 (18.9) | 27 (16.0) | 2,695 (17.7) | 9,842 (16.9) |
| White | 142 (33.2) | 107 (63.3) | 8,592 (56.6) | 40,217 (69.1) |
| **Ethnicity (%)** |  |  |  |  |
| Hispanic/Latino | 69 +/-5 (16.1) | 26 +/-5 (15.4) | 3,064 (20.2) | 11,416 (19.6) |
| Not Hispanic/Latino | 354 +/-5 (82.7) | 128 +/-5 (75.7) | 11,869 (78.1) | 45,119 (77.5) |
| Unknown | <20 | <20 | 260 (1.7) | 1,647 (2.8) |
| **Age group (%)** |  |  |  |  |
| 18-25 | <20 | <20 | 790 (5.2) | 7,573 (13.0) |
| 26-45 | 86 +/-5 (20.1) | 75 (44.4) | 3,824 (25.2) | 22,732 (39.1) |
| 46-65 | 188 +/-5 (43.9) | 69 (40.8) | 5,249 (34.5) | 19,015 (32.7) |
| 66+ | 147 +/-5 (34.3) | <20 | 5,330 (35.1) | 8,862 (15.2) |
| **Pre-COVID comorbidities (%)** |  |  |  |  |
| Diabetes | 86 (20.1) | <20 | 2,412 (15.9) | 4,842 (8.3) |
| Chronic kidney disease | 70 (16.4) | <20 | 1,721 (11.3) | 2,272 (3.9) |
| Congestive heart failure | 48 (11.2) | <20 | 960 (6.3) | 1,133 (1.9) |
| Chronic pulmonary disease | 45 (10.5) | 29 (17.2) | 1,415 (9.3) | 3,698 (6.4) |

**Table 2.** Demographic breakdown of potential long-COVID patients in the N3C cohort. We ran the trained “all patients’’ model on our base population of COVID-19 patients within the N3C Enclave (n = 846,981) with a predicted probability threshold set at 0.45 to emphasize recall. Age, sex, race, and ethnicity breakdowns of these patients are shown here for all patients as well as broken down by +/- potential long-COVID.

|  | All patients<br>n = 846,981 | Potential<br>long-COVID<br>n = 100,263 | Not potential<br>long-COVID<br>n = 746,718 |
| --- | --- | --- | --- |
| **Sex (%)** |  |  |  |
| Female | 500,531 (59.1) | 60,391 (60.2) | 440,140 (58.9) |
| Male | 340,984 (40.3) | 39,439 (39.3) | 301,545 (40.4) |
| Unknown | 5,466 (0.7) | 433 (0.4) | 5,033 (0.7) |
| **Race (%)** |  |  |  |
| Asian | 22,359 (2.6) | 2,339 (2.3) | 20,020 (2.7) |
| Black | 134,916 (15.9) | 17,611 (17.6) | 117,305 (15.7) |
| Native Haw./Pac Islander | 2,461 (0.3) | 279 (0.3) | 2,182 (0.3) |
| Other | 14,179 (1.7) | 1,668 (1.7) | 12,511 (1.7) |
| Unknown | 122,788 (14.5) | 13,075 (13.0) | 109,713 (14.7) |
| White | 550,278 (65.0) | 65,291 (65.1) | 484,987 (64.9) |
| **Ethnicity (%)** |  |  |  |
| Hispanic/Latino | 125,054 (14.8) | 15,658 (15.6) | 109,396 (14.7) |
| Not Hispanic/Latino | 659,079 (77.8) | 78,339 (78.1) | 580,740 (77.8) |
| Unknown | 62,848 (7.4) | 6,266 (6.2) | 56,582 (7.6) |
| **Age group (%)** |  |  |  |
| 18-25 | 100,232 (11.8) | 7,532 (7.5) | 92,700 (12.4) |
| 26-45 | 279,525 (33.0) | 29,940 (29.9) | 249,585 (33.4) |
| 46-65 | 292,445 (34.5) | 40,154 (40.0) | 252,291 (33.8) |
| 66+ | 174,779 (20.6) | 22,637 (22.6) | 152,142 (20.3) |

Each model was run against this three-site population; the performance metrics including receiver operating characteristic (ROC) curves are shown in Figure 2. For the purpose of calculating these metrics, long-COVID clinic patients are considered as actual positives; patients from the three sites who have not visited the specialty clinic are counted as true negatives. Patients labeled by the model as potential long-COVID patients should therefore be interpreted as “patients likely to be referred to or seek care at a long-COVID specialty clinic;” a proxy for long-COVID diagnosis in the current absence of a definition.

**Figure 2.**
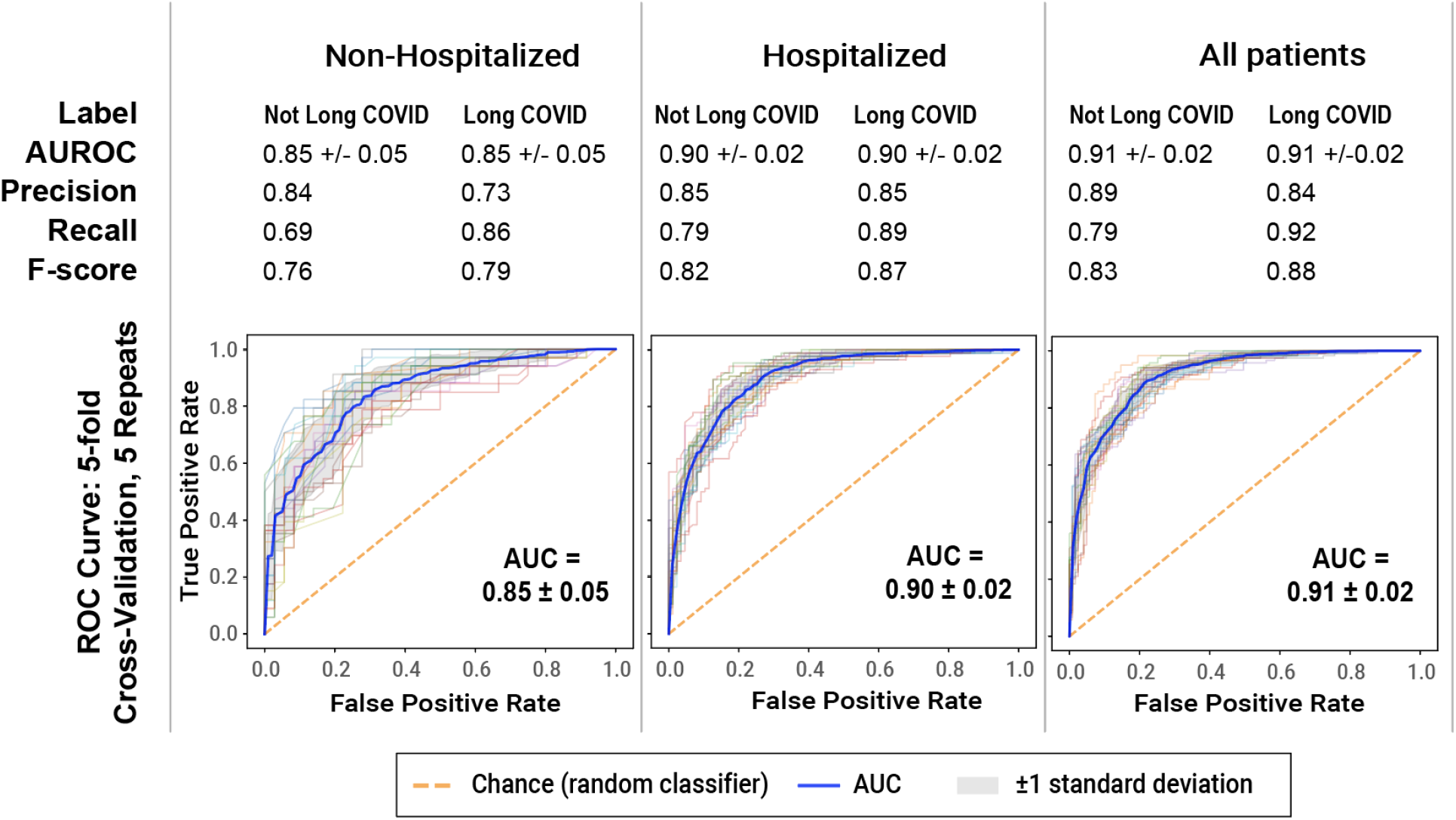
ML model performance in identifying potential long-COVID patients. Shown are receiver operating characteristic (ROC) curves, identifying the ability of each model (non-hospitalized, hospitalized, and all patients) to classify long-COVID patients as the discrimination threshold is varied. To emphasize recall of potential long-COVID patients, all models use a predicted probability threshold of 0.45 to generate the precision, recall, and F-score. The threshold can be adjusted to emphasize precision or recall, depending on the use case. All three models demonstrate robust performance.

Figure 3 shows the top 20 most important features (as determined using SHAP values) for each model; the top 50 most important features for each model are available in Supplemental Table 2a-c. Figure 4 shows the aggregate feature importance and univariate odds ratios for each model.

**Figure 3.**
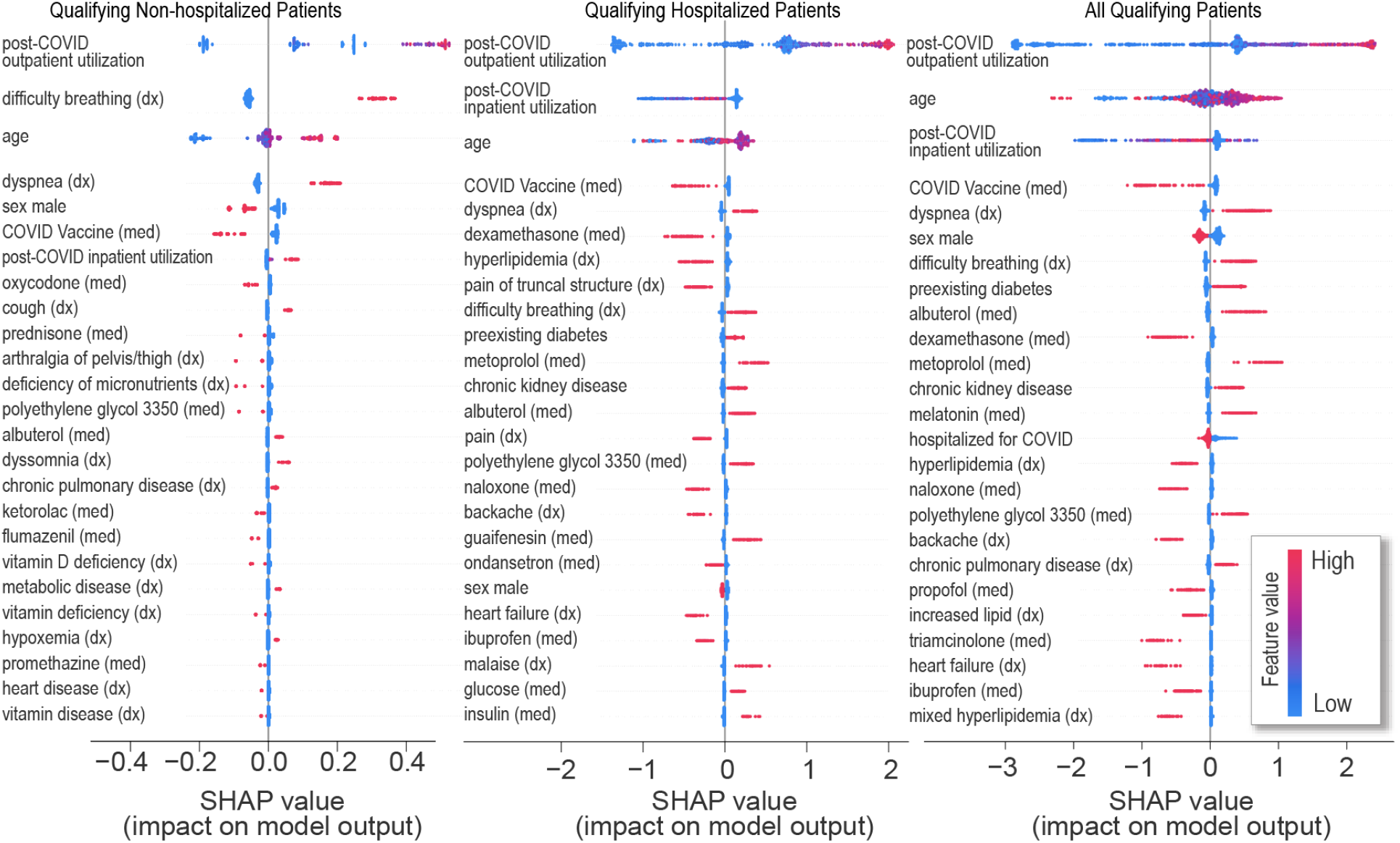
Most important model features for predicting visit to a long-COVID clinic. The top 20 features for each model are shown. Each point on the plot is a Shapley (importance) value for a single patient. The color of each point represents the magnitude and direction of the value of that feature for that patient. The point’s position on the horizontal axis represents the importance and direction of that feature for the prediction for that patient. Some features are important predictors in all models (e.g. outpatient utilization, dyspnea, COVID-19 vaccine), whereas others are specific to one or two of the models (e.g. dyssomnia, dexamethasone). Conditions labelled “chronic” were associated with patients prior to their COVID-19 index.

**Figure 4.**
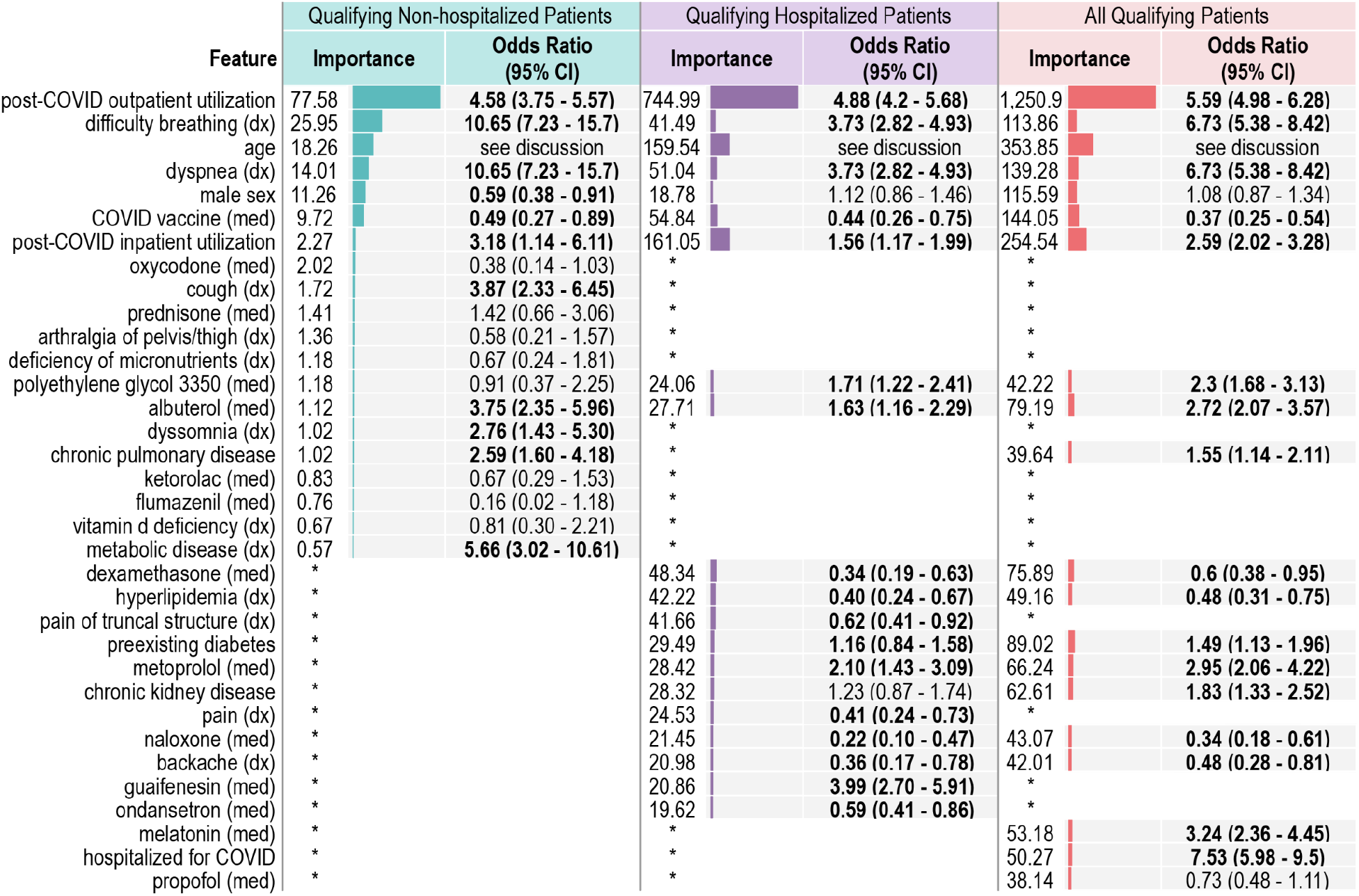
Many important features clearly differentiate potential long-COVID patients from non-long-COVID patients. Shown are the relative feature importance and univariate odds ratios for the top features (union of the 20 most important features) in each model. Odds ratios exclude age, which has a non-linear relationship with long-COVID. Regardless of importance, some features are significantly more prominent in the long-COVID clinic population, while others are more prominent in the non-long-COVID clinic population. Asterisks denote that the feature was not in the top 20 features for the model in that column. Conditions labelled “chronic” were associated with patients prior to their COVID-19 index.

## Interpretation

### Important model features

To avoid influencing the model with prior assumptions, we took a light-touch approach to feature selection, performing as little manual curation of features as possible prior to training and testing our models. Because of this approach, the reasons that a given feature may be important to one or more of the models is not always obvious. However, review by clinical experts of the features shown in Figures 3 and 4 and Supplementary Table 2a-c revealed a number of possible themes, described here.

### Post-COVID respiratory symptoms and associated treatments

These features are commonly reported for long-COVID patients.^7,9,22^ A confounding factor that prioritizes these features may be that the long-COVID clinics at two of the three sites contributing long-COVID clinic patients are based in the pulmonary department. However, given that SARS-CoV-2 is a primary respiratory virus, it is not surprising that long-term respiratory symptoms were observed. Similar long-term respiratory symptomatology is well-described with respiratory viral syndromes including those from severe acute respiratory syndrome, respiratory syncytial virus, influenza, and COVID-19.^23,24^ The high proportion of albuterol use and use of inhaled steroids is consistent with the expected high prevalence of post-viral reactive airways disease. Example features include dyspnea/difficulty breathing, cough, albuterol, guaifenesin, and hypoxemia.

### Non-respiratory symptoms widely reported as part of long-COVID and associated treatments

Sleep disorders, anxiety, malaise, chest pain, and constipation have all been reported as symptoms of long-COVID, and are included in WHO’s recent case definition.^10^ The example features in this group include both symptoms and mitigating treatments. Example features include dyssomnia, chest pain, malaise; and treatments with lorazepam, melatonin, and polyethylene glycol 3350.

### Preexisting risk factors for greater acute COVID severity

Some known risk factors for acute COVID-19 and severity are associated with long-COVID. This includes chronic conditions like diabetes, chronic kidney disease, and chronic pulmonary disease, which predispose patients at increased risk for worsened symptoms.^25^

### Proxies for hospitalization

Features that are representative of standard hospital admission orders are likely contributing to the model as proxies for hospitalization in general rather than being individually meaningful. These features tend to feature more prominently in the non-long-COVID group, suggesting that the model is (correctly) differentiating between acute illness requiring hospitalization and long-COVID. Example features include the use of glucose, ketorolac, propofol, and naloxone.

As shown in Figures 3 and 4 and Supplementary Table 2a-c, while there is considerable overlap between the most important features across the three models, there are also distinct differences. Notable differences include the high importance of dexamethasone in the hospitalized model, which decreased the likelihood of being labeled a potential long-COVID patient. Dexamethasone is not present in the top 50 features of the non-hospitalized model.

Similarly, cough and dyssomnia, which increased the likelihood of being labeled a potential long-COVID patient, are important features in the non-hospitalized model, but do not appear in the hospitalized model. COVID-19 vaccination after acute disease, consistently an important feature in all three models, decreased the likelihood of patients being labeled potential long-COVID. This result is noteworthy, and indicates that not only does vaccination against COVID-19 protect against hospitalization and death, but that it may also protect against long-COVID.

Rates of outpatient and inpatient utilization are clearly important features in all three models. This can be interpreted in a number of ways—patients who continue to feel unwell long after acute COVID-19 may be more likely to visit their providers repeatedly than those patients who fully recover. Because diagnosing and treating the heterogenous symptoms of long-COVID is an ongoing challenge, these patients may be referred to one or more specialists, increasing their utilization even more.

ML models do not consider each feature individually; rather, complex relationships between features can greatly influence classification. Each patient has their own path through the model, based on the data available about that individual. Figure 5 illustrates the path taken by three hypothetical patients through each of our three models, respectively. Information of this type is useful to make the outcomes of the ML models interpretable.

**Figure 5.**
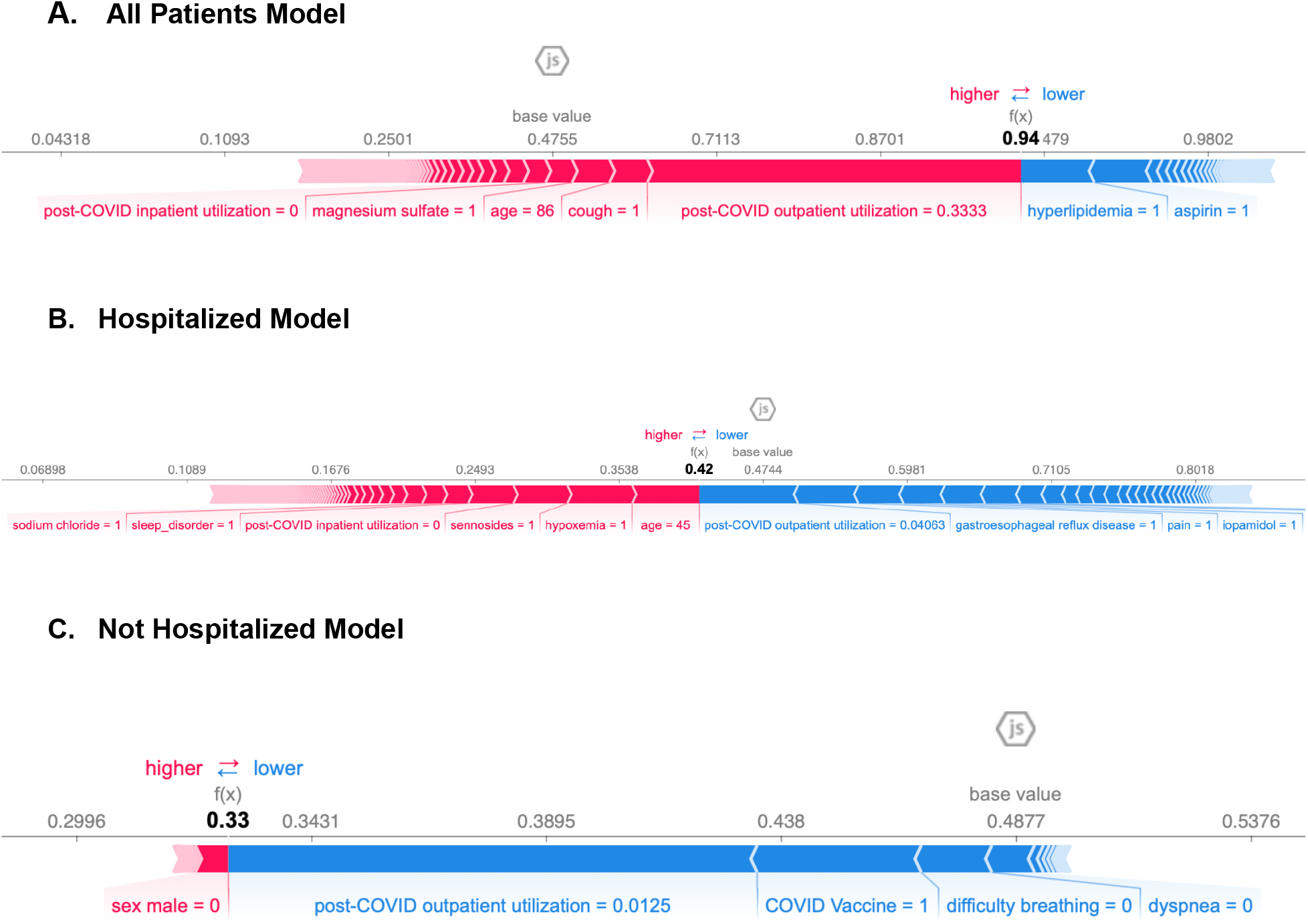
Example paths taken by the ML models to classify potential long-COVID patients. These Force plots illustrate the contribution of individual features to the final predicted probability of long-COVID generated by each of the three models for individual synthetic patients. Features colored red increase the probability of a long-COVID prediction, while features colored blue decrease that probability. The width of the bar for a given feature is proportional to the impact that feature has on the prediction for that patient. The final predicted probability is shown in bold text. 6A shows the “all patients” model, 6B the hospitalized model, and 6C the non-hospitalized model.

### Use of EHR data for long-COVID phenotyping

Though it contains rich clinical features, EHR data is also a proxy for healthcare utilization and can be interpreted through that lens. Diagnoses coded in the EHR are not representative of the *whole* patient, but rather are focused on the specific reasons the patient has come to the clinic or hospital on that day. Hospitalized and non-hospitalized patients are likely quite clinically different—but inpatient and outpatient coding practices are also different. Some of the differences between the hospitalized and non-hospitalized models may be an artifact of EHR data itself, and thus divergences between the two models are likely a result of both factors.

However, even as a proxy for utilization, EHR data is particularly well-suited to the task of cohort definition by way of computable phenotyping, especially when the end goal is study recruitment. Balancing inclusion and exclusion criteria is critically important to sufficiently define as homogeneous a population as possible, while still retaining key diversity and other recruitment goals (such as age distribution). While there are other methods of identifying potential study participants, a computable phenotype allows us to efficiently narrow the recruitment pool down from “everyone” to “patients likely to qualify”— easily eliminating large numbers of patients that do not qualify, and ascertaining patients that may elude human curation.

There are additional advantages to utilizing EHR data to identify long-COVID patients. With no single definition and no gold standard to compare with, the EHR allows us to define proxies for a condition and select on those—in this case, a patient’s visit to a long-COVID specialty clinic. However, rather than settling for a very restrictive inclusion criterion of “must have at least one visit to a long-COVID speciality clinic,” our ML models allow us to decouple patients’ utilization patterns from the clinic visit, meaning that we can use the models to identify similar patients who may not have access to a long-COVID clinic. Moreover, institutions without a long-COVID specialty clinic will be able to make use of the model for cohort identification within their own data. While using a proxy all but ensures that not all of the patients identified by the model have long-COVID, the resulting utilization patterns allow us to make some educated assumptions.

EHR data is skewed toward patients who make more use of health care systems, and is further skewed toward high utilizers, sicker patients, and inpatients. When we train models on N3C’s EHR data, it is essential to acknowledge whose data is less likely to be represented--uninsured patients, patients with limited access to or ability to pay for care, or patients seeking care at small practices or community hospitals with limited data exchange capabilities. Thus, while we believe our models are highly useful for identification and recruitment of long-COVID study participants, they should be used as one of several complementary recruitment methods to enable the widest possible reach and the most representative participants in research.

We did not include race and ethnicity as model features, as we did not believe our three-site sample of long-COVID clinic patients to be appropriately representative. As more long-COVID patients and training data are available over time, we will have the ability to balance the cohort based on demographics and, critically, to carefully account for race and ethnicity in future iterations of the model.

Beyond identifying cohorts for research studies, the models presented here can be applied in a variety of applications and could be enhanced in a number of ways. Specifically, it will undoubtedly be necessary to utilize a large sample size of long-COVID patients to validate hypotheses relating to social determinants of health and demographics, comorbidities, and treatment implications. It is also critical to understand the relationship between acute COVID-19 severity and specific long-COVID signs and symptoms and their longitudinal progression, as well as the influence of vaccination in such trajectories. The inclusion of additional data resources such as patient surveys, physiological data (e.g. sleep data), and wearables may further suggest EHR features that allow stratification to better inform mechanistic studies and therapy selection. Finally, it is plausible that long-COVID will not ultimately have a single definition, and may be better described as a set of related conditions with their own symptoms, trajectories, and treatments. Thus, as larger cohorts of long-COVID patients are established, future research should identify sub-phenotypes of long-COVID by clustering long-COVID patients with similar EHR data “fingerprints.” Future iterations of our models could discern among these clusters given N3C’s large sample size and recurring data feeds.

## Data Availability

The N3C data transfer to NCATS is performed under a Johns Hopkins University Reliance Protocol # IRB00249128 or individual site agreements with NIH. The N3C Data Enclave is managed under the authority of the NIH; information can be found at ncats.nih.gov/n3c/resources. Enclave data is protected, and can be accessed for COVID-related research with an approved (1) IRB protocol and (2) Data Use Request (DUR). A detailed accounting of data protections and access tiers is found in [1]. Enclave and data access instructions can be found at https://covid.cd2h.org/for-researchers; all code used to produce the analyses in this manuscript is available within the N3C Enclave to users with valid login credentials to support reproducibility.

## Acknowledgements

This research was possible because of the patients whose information is included within the data from participating organizations (covid.cd2h.org/dtas) and the organizations and scientists (covid.cd2h.org/duas) who have contributed to the on-going development of this community resource (cite this https://doi.org/10.1093/jamia/ocaa196).

## Individual Acknowledgements

We gratefully acknowledge the following contributors: David A. Eichmann, Justin Guinney, Warren A. Kibbe, Hongfang Liu, Philip R.O. Payne, Peter N. Robinson, Joel H. Saltz, Heidi Spratt, Justin Starren, Christine Suver, Adam B. Wilcox, Andrew E. Williams, Chunlei Wu, Davera Gabriel, Stephanie S. Hong, Harold P. Lehmann, Matvey B. Palchuk, Xiaohan Tanner Zhang, Richard L. Zhu, Benjamin Amor, Mark M. Bissell, Marshall Clark, Kristin Kostka, Adam M. Lee, Robert T. Miller, Matvey B. Palchuk, Kellie M. Walters, Karthik Natarajan, Shyam Visweswaran, Yooree Chae, Thomas Dillon, Patricia A. Francis, Rafael Fuentes, Alexis Graves, Shawn T. O’Neil, Usman Sheikh, Elizabeth Zampino, Christopher P. Austin, Leonie Misquitta, Xinzhi Zhang, Samuel Bozzette, Mariam Deacy, Nicole Garbarini, Michael G. Kurilla, Sam G. Michael, Joni L. Rutter, Meredith Temple-O’Connor, Benjamin Amor, Mark M. Bissell, Katie Rebecca Bradwell, Mary Morrison Saltz, Christine Suver, Mary Emmett, Victor Garcia, Jeremy Richard Harper, Wenndy Hernandez, Farrukh M Koraishy, Federico Mariona, Amit Saha, Satyanarayana Vedula. We acknowledge the Patient-Led Research Collaborative for their partnership, specifically Lisa McCorkell, Gina S. Assaf, Hannah E. Davis, and Hannah Wei.

## Data Partners with Released Data

Stony Brook University — U24TR002306 • University of Oklahoma Health Sciences Center — U54GM104938: Oklahoma Clinical and Translational Science Institute (OCTSI) • West Virginia University — U54GM104942: West Virginia Clinical and Translational Science Institute (WVCTSI) • University of Mississippi Medical Center — U54GM115428: Mississippi Center for Clinical and Translational Research (CCTR) • University of Nebraska Medical Center — U54GM115458: Great Plains IDeA-Clinical & Translational Research • Maine Medical Center — U54GM115516: Northern New England Clinical & Translational Research (NNE-CTR) Network • Wake Forest University Health Sciences — UL1TR001420: Wake Forest Clinical and Translational Science Institute • Northwestern University at Chicago — UL1TR001422: Northwestern University Clinical and Translational Science Institute (NUCATS) • University of Cincinnati — UL1TR001425: Center for Clinical and Translational Science and Training • The University of Texas Medical Branch at Galveston — UL1TR001439: The Institute for Translational Sciences • Medical University of South Carolina — UL1TR001450: South Carolina Clinical & Translational Research Institute (SCTR) • University of Massachusetts Medical School Worcester — UL1TR001453: The UMass Center for Clinical and Translational Science (UMCCTS) • University of Southern California — UL1TR001855: The Southern California Clinical and Translational Science Institute (SC CTSI) • Columbia University Irving Medical Center — UL1TR001873: Irving Institute for Clinical and Translational Research • George Washington Children’s Research Institute — UL1TR001876: Clinical and Translational Science Institute at Children’s National (CTSA-CN) • University of Kentucky — UL1TR001998: UK Center for Clinical and Translational Science • University of Rochester — UL1TR002001: UR Clinical & Translational Science Institute • University of Illinois at Chicago — UL1TR002003: UIC Center for Clinical and Translational Science • Penn State Health Milton S. Hershey Medical Center — UL1TR002014: Penn State Clinical and Translational Science Institute • The University of Michigan at Ann Arbor — UL1TR002240: Michigan Institute for Clinical and Health Research • Vanderbilt University Medical Center — UL1TR002243: Vanderbilt Institute for Clinical and Translational Research • University of Washington — UL1TR002319: Institute of Translational Health Sciences • Washington University in St. Louis — UL1TR002345: Institute of Clinical and Translational Sciences • Oregon Health & Science University — UL1TR002369: Oregon Clinical and Translational Research Institute • University of Wisconsin-Madison — UL1TR002373: UW Institute for Clinical and Translational Research • Rush University Medical Center — UL1TR002389: The Institute for Translational Medicine (ITM) • The University of Chicago — UL1TR002389: The Institute for Translational Medicine (ITM) • University of North Carolina at Chapel Hill — UL1TR002489: North Carolina Translational and Clinical Science Institute • University of Minnesota — UL1TR002494: Clinical and Translational Science Institute Children’s Hospital Colorado — UL1TR002535: Colorado Clinical and Translational Sciences Institute • The University of Iowa — UL1TR002537: Institute for Clinical and Translational Science • The University of Utah — UL1TR002538: Uhealth Center for Clinical and Translational Science • Tufts Medical Center — UL1TR002544: Tufts Clinical and Translational Science Institute • Duke University — UL1TR002553: Duke Clinical and Translational Science Institute • Virginia Commonwealth University — UL1TR002649: C. Kenneth and Dianne Wright Center for Clinical and Translational Research • The Ohio State University — UL1TR002733: Center for Clinical and Translational Science • The University of Miami Leonard M. Miller School of Medicine — UL1TR002736: University of Miami Clinical and Translational Science Institute • University of Virginia — UL1TR003015: iTHRIV Integrated Translational health Research Institute of Virginia • Carilion Clinic — UL1TR003015: iTHRIV Integrated Translational health Research Institute of Virginia • University of Alabama at Birmingham — UL1TR003096: Center for Clinical and Translational Science • Johns Hopkins University — UL1TR003098: Johns Hopkins Institute for Clinical and Translational Research • University of Arkansas for Medical Sciences — UL1TR003107: UAMS Translational Research Institute • Nemours — U54GM104941: Delaware CTR ACCEL Program • University Medical Center New Orleans — U54GM104940: Louisiana Clinical and Translational Science (LA CaTS) Center • University of Colorado Denver, Anschutz Medical Campus — UL1TR002535: Colorado Clinical and Translational Sciences Institute • Mayo Clinic Rochester — UL1TR002377: Mayo Clinic Center for Clinical and Translational Science (CCaTS) • Tulane University — UL1TR003096: Center for Clinical and Translational Science • Loyola University Medical Center — UL1TR002389: The Institute for Translational Medicine (ITM) • Advocate Health Care Network — UL1TR002389: The Institute for Translational Medicine (ITM) • OCHIN — INV-018455: Bill and Melinda Gates Foundation grant to Sage Bionetworks • The University of Texas Health Science Center at Houston — UL1TR003167: Center for Clinical and Translational Sciences (CCTS) • Weill Medical College of Cornell University — UL1TR002384: Weill Cornell Medicine Clinical and Translational Science Center • Montefiore Medical Center — UL1TR002556: Institute for Clinical and Translational Research at Einstein and Montefiore • Regenstrief Institute — UL1TR002529: Indiana Clinical and Translational Science Institute • Boston University Medical Campus — UL1TR001430: Boston University Clinical and Translational Science Institute • Aurora Health Care — UL1TR002373: Wisconsin Network For Health Research • Brown University — U54GM115677: Advance Clinical Translational Research (Advance-CTR) • Rutgers, The State University of New Jersey — UL1TR003017: New Jersey Alliance for Clinical and Translational Science • Loyola University Chicago — UL1TR002389: The Institute for Translational Medicine (ITM) • UL1TR001445: Langone Health’s Clinical and Translational Science Institute • University of Kansas Medical Center — UL1TR002366: Frontiers: University of Kansas Clinical and Translational Science Institute • Massachusetts General Brigham — UL1TR002541: Harvard Catalyst • University of California, Irvine — UL1TR001414: The UC Irvine Institute for Clinical and Translational Science (ICTS) • University of California, San Diego — UL1TR001442: Altman Clinical and Translational Research Institute • University of California, Davis — UL1TR001860: UCDavis Health Clinical and Translational Science Center • University of California, San Francisco — UL1TR001872: UCSF Clinical and Translational Science Institute • University of California, Los Angeles — UL1TR001881: UCLA Clinical Translational Science Institute •

## Additional Data Partners Who Have Signed a DTA and Whose Data Release is Pending

The Scripps Research Institute — UL1TR002550: Scripps Research Translational Institute • University of Texas Health Science Center at San Antonio — UL1TR002645: Institute for Integration of Medicine and Science • NorthShore University HealthSystem — UL1TR002389: The Institute for Translational Medicine (ITM) • Yale New Haven Hospital — UL1TR001863: Yale Center for Clinical Investigation • Emory University — UL1TR002378: Georgia Clinical and Translational Science Alliance • Medical College of Wisconsin — UL1TR001436: Clinical and Translational Science Institute of Southeast Wisconsin • University of New Mexico Health Sciences Center — UL1TR001449: University of New Mexico Clinical and Translational Science Center • George Washington University — UL1TR001876: Clinical and Translational Science Institute at Children’s National (CTSA-CN) • Stanford University — UL1TR003142: Spectrum: The Stanford Center for Clinical and Translational Research and Education • Cincinnati Children’s Hospital Medical Center — UL1TR001425: Center for Clinical and Translational Science and Training • The State University of New York at Buffalo — UL1TR001412: Clinical and Translational Science Institute • Children’s Hospital of Philadelphia — UL1TR001878: Institute for Translational Medicine and Therapeutics • Icahn School of Medicine at Mount Sinai UL1TR001433: ConduITS Institute for Translational Sciences • Ochsner Medical Center — U54GM104940: Louisiana Clinical and Translational Science (LA CaTS) Center • HonorHealth None (Voluntary) • University of Vermont — U54GM115516: Northern New England Clinical & Translational Research (NNE-CTR) Network • Arkansas Children’s Hospital — UL1TR003107: UAMS Translational Research Institute

## Author Contributions

- data curation: ER Pfaff, AT Girvin, K Kostka, CG Chute, MA Haendel
- data integration: ER Pfaff, AT Girvin, MG Kahn, K Kostka, CG Chute
- data quality assurance: ER Pfaff, AT Girvin, MG Kahn, CG Chute
- N3C Phenotype definition: ER Pfaff, K Kostka, CG Chute
- clinical data model expertise: ER Pfaff, AT Girvin, TD Bennett, MG Kahn, K Kostka, CG Chute
- clinical subject matter expertise: TD Bennett, RR Deer, SE Jolley
- statistical analysis: AT Girvin, Abhishkek Bhatia, Jonathan P Dekermanjian,
- data visualization: AT Girvin, A Bhatia, JP Dekermanjian, JA McMurry, MA Haendel
- critical revision of the manuscript for important intellectual content: ER Pfaff, AT Girvin,TD Bennett, IM Brooks, RR Deer, SE Jolley, MG Kahn, JA McMurry, RA Moffitt, A Walden, K Kostka, CG Chute, MA Haendel
- manuscript drafting: ER Pfaff, AT Girvin, RR Deer, SE Jolley, JA McMurry, CG Chute, MA Haendel
- governance/regulatory oversight: JA McMurry, A Walden, CG Chute, MA Haendel

Authors ER Pfaff and AT Girvin have verified all underlying data for these analyses.

## Ethics and Regulatory

The N3C data transfer to NCATS is performed under a Johns Hopkins University Reliance Protocol # IRB00249128 or individual site agreements with NIH.

Use of the N3C data for this study is authorized under the following IRB Protocols:

| Site | IRB name | Exempted vs approved | Protocol number |
| --- | --- | --- | --- |
| University of Colorado | Colorado Multiple Institutional Review Board | approved | 21-2759 |
| Johns Hopkins University | Johns Hopkins Office of Human Subjects Research - Institutional Review Board | approved | IRB00249128 |
| University of North Carolina | University of North Carolina Chapel Hill Institutional Review Board | exempt | 21-0309 |
| Stony Brook University | Office of Research Compliance, Division of Human Subject Protections, Stony Brook University | exempt | IRB2021-00098 |

