## Supplemental Table 1 for "Who has long-COVID? A big data approach"

### Supplemental Figure 1: Cohort Selection Flow Diagram

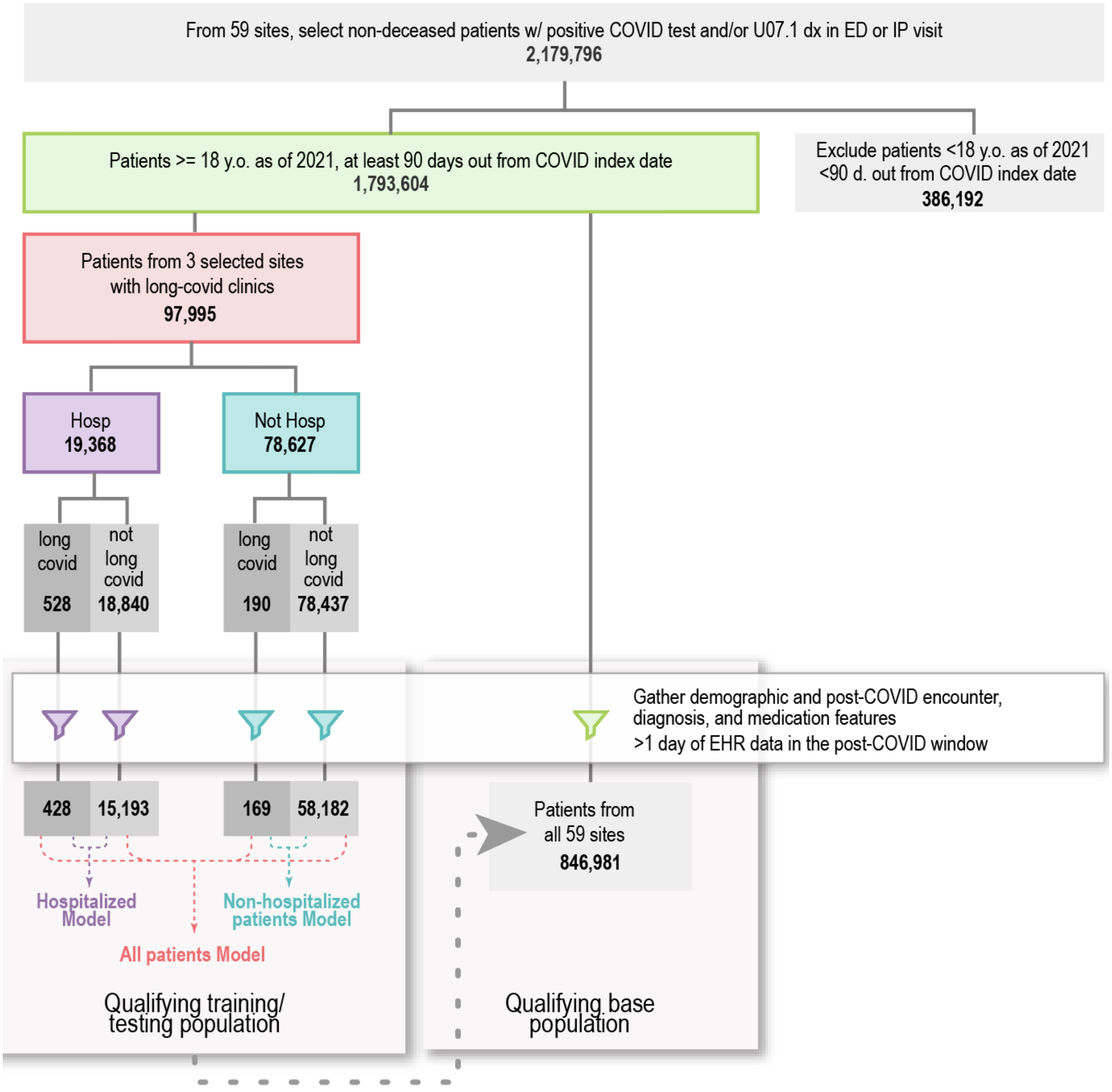

### Supplemental Methods: Feature Engineering

The feature tables used as inputs for each model are made up of patient age, sex, healthcare utilization metrics, pre-COVID-19 comorbidities, post-COVID-19 diagnoses, and post-COVID-19 medications. Curation methods for each of these domains are described here. All OMOP concept sets and feature engineering code described here are available by author request via the N3C Enclave, according to the N3C governance and regulatory policies.

#### Healthcare utilization metrics

In order to calculate a utilization metric for each patient, we first determine the number of days in each patient's post-COVID-19 "window." By our definition, this window begins 45 days after the patient's COVID-19 index date and ends 320 days later, for a total of 365 days after the COVID-19 index date. Thus, every patient has a window of 320 days (though the dates differ for each patient, based on their COVID-19 index dates). The exception to this is the long-COVID clinic patients used to train the model; their post-COVID-19 window ends on the day before their first long-COVID clinic visit date, as supplied by the three sites.

Outpatient utilization ratios for each patient are calculated as:

$$\frac{\text{(\# unique calendar days patient has an outpatient encounter in the post-COVID-19 window)}}{\text{(\# days in the patient's post-COVID-19 window)}}$$

Inpatient utilization ratios for each patient are calculated as:

$$\frac{\text{(sum of lengths of stay for all inpatient visits in the post-COVID-19 window)}}{\text{(\# days in the patient's post-COVID-19 window)}}$$

#### Pre-COVID-19 comorbidities

We added indicator variables to flag whether each patient had one or more of the following comorbidities *prior* to their COVID-19 index date: diabetes, chronic kidney disease, congestive heart failure, chronic pulmonary disease. A patient was flagged as having one of these comorbidities if they had a diagnosis code for that condition on two or more occasions (distinct calendar days) prior to their COVID-19 index date.

#### Post-COVID-19 diagnoses

The OMOP data model uses SNOMED CT as a standard vocabulary for diagnoses. SNOMED CT is multi-hierarchical, meaning that a single child concept can have many parent concepts. Here, we "rolled up" SNOMED-coded diagnoses to parent-level features for use in our ML models. Because the various levels of the SNOMED hierarchy are not consistent in granularity, automated roll up can lead to uninformative terms that are not useful model features (e.g. "disorder of body system"), and these uninformative concepts can appear as model features if enough of their child concepts have some importance in classification.

Informative terms that should be included based on clinical relevance are sometimes present at the same hierarchical level as these less informative terms. It is therefore not possible to roll up all SNOMED terms using a specific hierarchical level (i.e., roll up diagnoses to the fourth level down from the root). We therefore devised a computational approach to rolling up terms that lessens uninformative terms. A summary of the approach follows:

- Extract a dataset of all SNOMED CT child terms (i.e., the diagnosis terms used in the patient data) matched with two levels of parent terms (i.e., the child term's parent(s) and the parent(s)' parent(s)).
- Exclude the following parent concepts entirely, due to non-informativeness: "105721009 - General problem AND/OR complaint," '64572001 - Disease," "363296001 - Sequelae of disorders classified by disorder-system," "362977000 - Sequela," "58184002 - Recurrent disease," "55607006 - Problem," "2704003 - Acute disease," "27624003 - Chronic disease," "116223007 - Complication." Child conditions that roll up to these parent terms remain in the data, but will not roll up to one of these terms.

- Exclude parent terms that contain the words “right” or “left”, which enables all child terms that specify laterality to be rolled up to a non-lateral parent.
- Exclude parent terms that contain the words “finding,” “disorder of,” or “by site” (which are generally generic, low-information terms), *unless* removal of the parent term would leave a child term without a viable parent term.
- Use the remaining set of parent terms in place of the child terms in the model.

Despite this computational pruning, there is still some concept duplication within our ML model because closely related concepts (which may be equivalent by some definitions) can exist in different branches of the hierarchy (e.g. “dyspnea” and “difficulty breathing”). These are inherent caveats of using ontologies and can be addressed through manual curation for future studies.

For each patient, we only counted conditions that newly occurred or occurred in greater frequency in the post-COVID-19 period compared to the pre-COVID-19 period. Finally, before running the model, we limited diagnosis features to those that were associated with at least 1% of the patients in our three-site subset. We first ran the model without this restriction to ensure this would not affect model performance, and noted our results were the same. We thus opted to add this restriction in order to render our models less computationally intensive.

#### Post-COVID-19 medications

As with diagnoses, we rolled up medication records to parent-level concepts in order to collapse related terms into single features. We used the OMOP vocabulary to roll up all medication records to the “ingredient” level. As shown in the example below, these roll-ups enable us to combine multiple forms of the same drug into a single term for modelling purposes. Combination drugs separately roll up to each of their ingredients, also shown below.

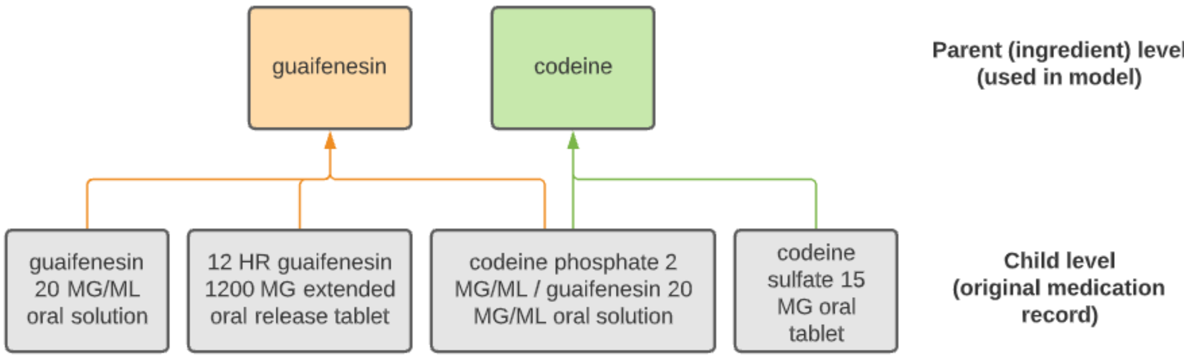


For each patient, we only counted medications that were newly prescribed in the post-COVID-19 period, with no records in the pre-COVID-19 period. Finally, before running the model, we limited medication features to those that were associated with at least 1% of the patients in our three-site subset. We first ran the model without this restriction to ensure this would not affect model performance, and noted our results were the same. We thus opted to add this restriction in order to render our models less computationally intensive.

### Supplemental Table 1: Training and Test Set Patient Counts

| | | Training | | Testing | |
|---|---|---|---|---|---|
| **Model** | **Patient subset** | **Long-COVID Clinic Patients** | **Not Long-COVID** | **Long-COVID Clinic Patients** | **Not Long-COVID** |
| All patients | Hospitalized | 327 | 360 | 101 | 76 |
| | Not hospitalized | 125 | 134 | 44 | 45 |
| Hospitalized | Hospitalized | 327 | 360 | 101 | 76 |
| Not-Hospitalized | Not hospitalized | 125 | 134 | 44 | 45 |

### Supplemental Table 2: Top 50 features in each model

The importance score for each feature is the sum of the importance of that feature for each patient. Mean values are the mean value of each feature across each group. All diagnosis and medication features are binary (1=yes, 0=no). For diagnoses, “yes” means that the patient had a greater number of occurrences of that diagnosis code in their post-COVID-19 window than their pre-COVID-19 window. For medications, “yes” means that the patient had one or more prescriptions for that medication in their post-COVID-19 window, and no record of that medication in their pre-COVID-19 window. Post-COVID-19 outpatient and inpatient utilization and age are continuous variables.

**Table 2a. Top 50 all-patients model features, ranked by importance (SHAP value)**.

| | | Mean Values | |
|---|---|---|---|
| **Feature** | **Importance** | **Not Long-COVID** | **Predicted Long-COVID** |
| post-COVID-19 outpatient utilization | 1,250.91 | 0.03 | 0.09 |
| age | 353.85 | 52.04 | 55.27 |
| post-COVID-19 inpatient utilization | 254.54 | 0.01 | 0.01 |
| COVID-19 vaccine (med) | 144.05 | 0.15 | 0.05 |
| dyspnea (dx) | 139.28 | 0.08 | 0.21 |
| male sex | 115.59 | 0.45 | 0.38 |
| difficulty breathing (dx) | 113.86 | 0.08 | 0.21 |
| preexisting diabetes | 89.02 | 0.12 | 0.17 |
| albuterol (med) | 79.19 | 0.07 | 0.12 |
| dexamethasone (med) | 75.89 | 0.11 | 0.04 |
| metoprolol (med) | 66.24 | 0.03 | 0.08 |
| preexisting chronic kidney disease | 62.61 | 0.08 | 0.12 |
| melatonin (med) | 53.18 | 0.06 | 0.10 |
| hospitalized for COVID-19 | 50.27 | 0.73 | 0.72 |
| hyperlipidemia (dx) | 49.16 | 0.11 | 0.04 |
| naloxone (med) | 43.07 | 0.08 | 0.02 |
| polyethylene glycol 3350 (med) | 42.22 | 0.05 | 0.10 |
| backache (dx) | 42.01 | 0.06 | 0.02 |
| preexisting chronic pulmonary disease | 39.64 | 0.08 | 0.12 |
| propofol (med) | 38.14 | 0.09 | 0.04 |
| increased lipids (dx) | 34.36 | 0.10 | 0.04 |
| triamcinolone (med) | 34.15 | 0.04 | 0.02 |
| heart failure (dx) | 30.77 | 0.04 | 0.01 |
| ibuprofen (med) | 30.04 | 0.07 | 0.03 |
| mixed hyperlipidemia (dx) | 27.28 | 0.04 | 0.01 |
| sennosides USP (med) | 24.99 | 0.06 | 0.09 |
| low back pain (dx) | 24.87 | 0.05 | 0.01 |
| nausea (dx) | 24.17 | 0.04 | 0.02 |
| cough (dx) | 23.62 | 0.04 | 0.06 |
| preexisting congestive heart failure | 22.66 | 0.06 | 0.08 |
| malaise (dx) | 22.37 | 0.01 | 0.05 |
| guaifenesin (med) | 20.25 | 0.03 | 0.07 |
| salmeterol (med) | 19.52 | 0.01 | 0.03 |
| aspirin (med) | 18.99 | 0.05 | 0.04 |
| soft tissue lesion (dx) | 18.37 | 0.04 | 0.02 |
| pain of truncal structure (dx) | 18.32 | 0.12 | 0.08 |
| pain (dx) | 17.85 | 0.09 | 0.04 |
| clinical finding (dx) | 17.41 | 0.02 | 0.04 |
| abdominal pain (dx) | 16.80 | 0.05 | 0.01 |
| acute respiratory disease (dx) | 16.74 | 0.05 | 0.03 |
| joint pain (dx) | 15.65 | 0.04 | 0.01 |
| renal failure syndrome (dx) | 15.60 | 0.04 | 0.01 |
| dyssomnia (dx) | 15.07 | 0.02 | 0.03 |
| amoxicillin (med) | 14.72 | 0.04 | 0.02 |
| hydralazine (med) | 14.29 | 0.03 | 0.03 |
| asthenia (dx) | 14.03 | 0.04 | 0.03 |
| diphenhydramine (med) | 13.59 | 0.09 | 0.04 |
| chest pain (dx) | 13.41 | 0.05 | 0.07 |
| tramadol (med) | 13.15 | 0.06 | 0.02 |
| hypertensive disorder (dx) | 13.12 | 0.12 | 0.08 |

**Table 2b. Top 50 non-hospitalized model features, ranked by importance (SHAP value)**

| | | Mean Values | |
|---|---|---|---|
| Feature | Importance | Not Long-COVID | Predicted Long-COVID |
| post-COVID-19 outpatient utilization | 77.58 | 0.02 | 0.07 |
| difficulty breathing (dx) | 25.95 | 0.04 | 0.29 |
| age | 18.26 | 41.94 | 48.46 |
| dyspnea (dx) | 14.01 | 0.04 | 0.29 |
| male sex | 11.26 | 0.37 | 0.24 |
| COVID-19 vaccine (med) | 9.72 | 0.22 | 0.08 |
| post-COVID-19 inpatient utilization | 2.27 | 0.00 | 0.00 |
| oxycodone (med) | 2.02 | 0.10 | 0.04 |
| cough (dx) | 1.72 | 0.01 | 0.10 |
| prednisone (med) | 1.41 | 0.04 | 0.04 |
| arthralgia of the pelvic region and thigh (dx) | 1.36 | 0.06 | 0.03 |
| deficiency of micronutrients (dx) | 1.18 | 0.01 | 0.03 |
| polyethylene glycol 3350 | 1.18 | 0.04 | 0.02 |
| albuterol (med) | 1.12 | 0.02 | 0.13 |
| dyssomnia (dx) | 1.02 | 0.01 | 0.06 |
| preexisting chronic pulmonary disease | 1.02 | 0.05 | 0.16 |
| ketorolac (med) | 0.83 | 0.07 | 0.04 |
| flumazenil (med) | 0.76 | 0.06 | 0.01 |
| vitamin D deficiency (dx) | 0.67 | 0.01 | 0.03 |
| metabolic disease (dx) | 0.57 | 0.00 | 0.08 |
| vitamin deficiency (dx) | 0.50 | 0.01 | 0.03 |
| hypoxemia (dx) | 0.47 | 0.00 | 0.08 |
| promethazine (med) | 0.34 | 0.05 | 0.01 |
| heart disease (dx) | 0.31 | 0.01 | 0.03 |
| vitamin disease (dx) | 0.29 | 0.01 | 0.03 |
| gabapentin (med) | 0.29 | 0.03 | 0.05 |
| clinical finding (dx) | 0.28 | 0.00 | 0.06 |
| diphenhydramine (med) | 0.28 | 0.07 | 0.04 |
| ibuprofen (med) | 0.21 | 0.07 | 0.03 |
| hydromorphone (med) | 0.19 | 0.07 | 0.02 |
| inflammation of specific body organs (dx) | 0.17 | 0.07 | 0.06 |
| benzocaine (med) | 0.17 | 0.06 | 0.02 |
| asthma (dx) | 0.17 | 0.04 | 0.05 |
| disorders of initiating and maintaining sleep (dx) | 0.17 | 0.01 | 0.06 |
| sleep disorder (dx) | 0.15 | 0.04 | 0.10 |
| hyperlipidemia (dx) | 0.14 | 0.04 | 0.04 |
| increased lipids (dx) | 0.14 | 0.04 | 0.03 |
| phenylephrine (med) | 0.12 | 0.06 | 0.02 |
| amoxicillin (med) | 0.12 | 0.05 | 0.02 |
| famotidine (med) | 0.09 | 0.04 | 0.02 |
| iohexol (med) | 0.09 | 0.00 | 0.04 |
| knee pain (dx) | 0.08 | 0.04 | 0.00 |
| chest pain (dx) | 0.08 | 0.06 | 0.11 |
| fentanyl (med) | 0.07 | 0.07 | 0.06 |
| pain of truncal structure (dx) | 0.07 | 0.10 | 0.12 |
| lactate (med) | 0.07 | 0.09 | 0.05 |
| muscle strain (dx) | 0.00 | 0.01 | 0.01 |
| musculoskeletal chest pain (dx) | 0.00 | 0.02 | 0.00 |
| muscle weakness (dx) | 0.00 | 0.00 | 0.02 |
| musculoskeletal finding (dx) | 0.00 | 0.00 | 0.04 |

**Table 2c. Top 50 hospitalized model features, ranked by importance (SHAP value)**

| | | Mean Values | |
|---|---|---|---|
| **Feature** | **Importance** | **Not Long-COVID** | **Predicted Long-COVID** |
| post-COVID-19 outpatient utilization | 744.99 | 0.03 | 0.10 |
| post-COVID-19 inpatient utilization | 161.05 | 0.01 | 0.01 |
| age | 159.54 | 55.80 | 57.87 |
| COVID-19 vaccine (med) | 54.84 | 0.12 | 0.04 |
| dyspnea (dx) | 51.04 | 0.09 | 0.18 |
| dexamethasone (med) | 48.34 | 0.12 | 0.03 |
| hyperlipidemia (dx) | 42.22 | 0.13 | 0.04 |
| pain of truncal structure (dx) | 41.66 | 0.12 | 0.06 |
| difficulty breathing (dx) | 41.49 | 0.09 | 0.18 |
| preexisting diabetes | 29.49 | 0.14 | 0.19 |
| metoprolol (med) | 28.42 | 0.04 | 0.09 |
| preexisting chronic kidney disease | 28.32 | 0.10 | 0.16 |
| albuterol (med) | 27.71 | 0.09 | 0.12 |
| pain (dx) | 24.53 | 0.10 | 0.03 |
| polyethylene glycol 3350 (med) | 24.06 | 0.05 | 0.12 |
| naloxone (med) | 21.45 | 0.08 | 0.01 |
| backache (dx) | 20.98 | 0.07 | 0.01 |
| guaifenesin (med) | 20.86 | 0.03 | 0.09 |
| ondansetron (med) | 19.62 | 0.15 | 0.09 |
| male sex | 18.78 | 0.48 | 0.44 |
| heart failure (dx) | 18.01 | 0.06 | 0.01 |
| ibuprofen (med) | 17.98 | 0.07 | 0.02 |
| malaise (dx) | 16.23 | 0.02 | 0.06 |
| glucose (med) | 15.82 | 0.08 | 0.09 |
| insulin (med) | 15.31 | 0.01 | 0.07 |
| sennosides USP (med) | 14.98 | 0.08 | 0.11 |
| gastroesophageal reflux disease without esophagitis (dx) | 14.93 | 0.08 | 0.03 |
| breathing related sleep disorder (dx) | 14.46 | 0.06 | 0.03 |
| diabetes mellitus without complication (dx) | 13.92 | 0.07 | 0.03 |
| nausea (dx) | 13.59 | 0.05 | 0.01 |
| clinical history and observation findings (dx) | 13.50 | 0.10 | 0.06 |
| chlorhexidine (med) | 12.92 | 0.01 | 0.04 |
| iohexol (med) | 12.61 | 0.05 | 0.08 |
| sodium chloride (med) | 12.24 | 0.14 | 0.12 |
| Am. Soc of anesthesiologists physical status classification (dx) | 11.97 | 0.08 | 0.04 |
| phenylephrine (med) | 11.71 | 0.06 | 0.03 |
| increased lipids (dx) | 11.54 | 0.12 | 0.04 |
| bupivacaine (med) | 10.77 | 0.05 | 0.00 |
| quetiapine (med) | 10.67 | 0.02 | 0.05 |
| iopamidol (med) | 10.46 | 0.07 | 0.02 |
| melatonin (med) | 10.46 | 0.08 | 0.13 |
| renal failure syndrome (dx) | 10.19 | 0.05 | 0.02 |
| heart disease (dx) | 10.04 | 0.07 | 0.04 |
| fentanyl (med) | 9.43 | 0.13 | 0.06 |
| diphenhydramine (med) | 9.32 | 0.10 | 0.04 |
| hypoxemia (dx) | 9.17 | 0.07 | 0.10 |
| triamcinolone (med) | 9.10 | 0.04 | 0.02 |
| deficiency of micronutrients (dx) | 8.74 | 0.04 | 0.03 |
| ergocalciferol (med) | 8.67 | 0.03 | 0.06 |
| obstructive sleep apnea syndrome (dx) | 8.57 | 0.06 | 0.02 |
